# A lateral flow test detecting SARS-CoV-2 neutralizing antibodies

**DOI:** 10.1101/2020.11.05.20222596

**Authors:** Nan Zhang, Shuo Chen, Jin V. Wu, Xinhai Yang, Jianfu J. Wang

**Affiliations:** Novodiax Inc. Hayward, CA 94587

## Abstract

It is of critical importance for COVID-19 survivors, vaccine recipients, and public to know whether they have developed neutralizing antibodies or immunity. Here, we describe a 15 minutes lateral flow test for rapid detection of neutralizing antibodies against SARS-CoV-2. All other currently available neutralization tests require hours or days to complete and have to be performed in a well-equipped laboratory. This lateral flow test is the first of its kind and will serve as a convenient diagnostic tool in management of COVID-19 disease.

## Main

Over 40 million confirmed COVID-19 cases and more than one million deaths caused by this disease have been reported as of October 2020.^1^ To combat this threat, more than 300 vaccines are under development^2^, with more than 30 of them in various clinical stages.^3^ The majority of these vaccines aim to target SARS-CoV-2’s spike (S) protein or the S protein’s receptor binding domain (RBD) in order to induce neutralizing antibodies (NAbs) that block the interaction of RBD with its receptor angiotensin-converting enzyme 2(ACE2) on host cells.^4^

Unlike other binding antibodies (BAbs), NAbs represent a type of humoral immunity capable of neutralizing or blocking viruses from entering host cells, preventing them from reproducing and causing severe damage. However, NAbs are not equally developed among the COVID-19 convalescent patients^5,6^ and IgG antibodies specific to RBD developed in patients with mild COVID-19 decay rapidly, with a half-life of approximately 36 days^7^. Multiple COVID-19 re-infection cases have been reported.^8-10^ Evaluation of SARS-CoV-2 NAbs is critical for both better understanding of immunity to COVID-19 and monitoring levels of protective immunity among the recipients of COVID-19 vaccines.

Various assay methods for SARS-CoV-2 NAbs are available. Conventional virus neutralization tests use live ACE2-expressing cells and live SARS-CoV-2^11^ and need to be performed in a biosafety level 3 laboratory. Several virus neutralization tests using pseudovirus^12-17^ have also been developed. These tests only need a biosafety level 2 facility and usually take several days to complete. A simpler and faster enzyme-linked immunosorbent assay (ELISA) that detects SARS-CoV-2 NAbs based on ACE2-RBD interaction has been recently reported.^18^ This new test mimics the virus-host interaction in an ELISA test which can be completed in a safety level 2 laboratory within a few hours. The lateral flow test (LFT) is the fastest and most convenient test among the popular immunoassays, which typically takes only 15 minutes to complete. It can be performed either in a professional laboratory or by an individual, thus if available should be a necessary complement to all available neutralizing tests. Here we report the development of the first COVID-19 NAb LFT, or Point of Care Test (POCT), along with several noteworthy insights that we’ve gleaned from its development.

Eighty patients’ plasmas were purchased from a Bio-Bank company, 50 of these were PCR positive for SARS-CoV-2 (named P1-P50) and 30 were PCR negative (N1-N30). Twenty normal plasmas (NP1-NP20) were randomly picked from an in-house collection dated between 2014 and 2016, before the COVID-19 pandemic, to establish negative baselines. With all of this in place, we developed an ELISA to evaluate all 100 plasmas. We then set up a pseudovirus based neutralization test and developed a Nab LFT. The latter test, the first of its kind to our knowledge, was compared with and evaluated against the other two popular assays.

Using the ELISA, we measured IgG and IgM BAbs specific to the spike (S) protein, envelop (E) protein and nucleocapsid (N) protein of SARS-CoV-2 for all 100 plasmas, and measured NAbs for the 80 patients’ plasmas. In line with previous findings, patients’ humoral immune response to SARS-CoV-2 varied widely, from completely negative to high titer of both BAbs and NAbs (Fig 1A-1C). Initial data analyses revealed that positive antibody response rates to N and E proteins were far lower than the response rates to S protein, and no patient responded to N or E protein without raising IgG or IgM against S protein. We thus focused only on S protein in the following development. Based on the relative sensitivity of the assay (patients’ plasmas vs the 20 normal plasmas), signals above average plus 2SD of that of the 20 normal plasmas at 1:200 dilution were chosen as the positive cutoff for both IgG and IgM. In general, PCR positive patients developed more detectable anti-S1 IgG than the PCR negative patients (78% versus 53%, p=0.0006) (Fig 1D-1F), and similar level of anti-S1 IgM (70% versus 66.7%, p=0.19) (Fig 1D-1F).

**Figure 1.**
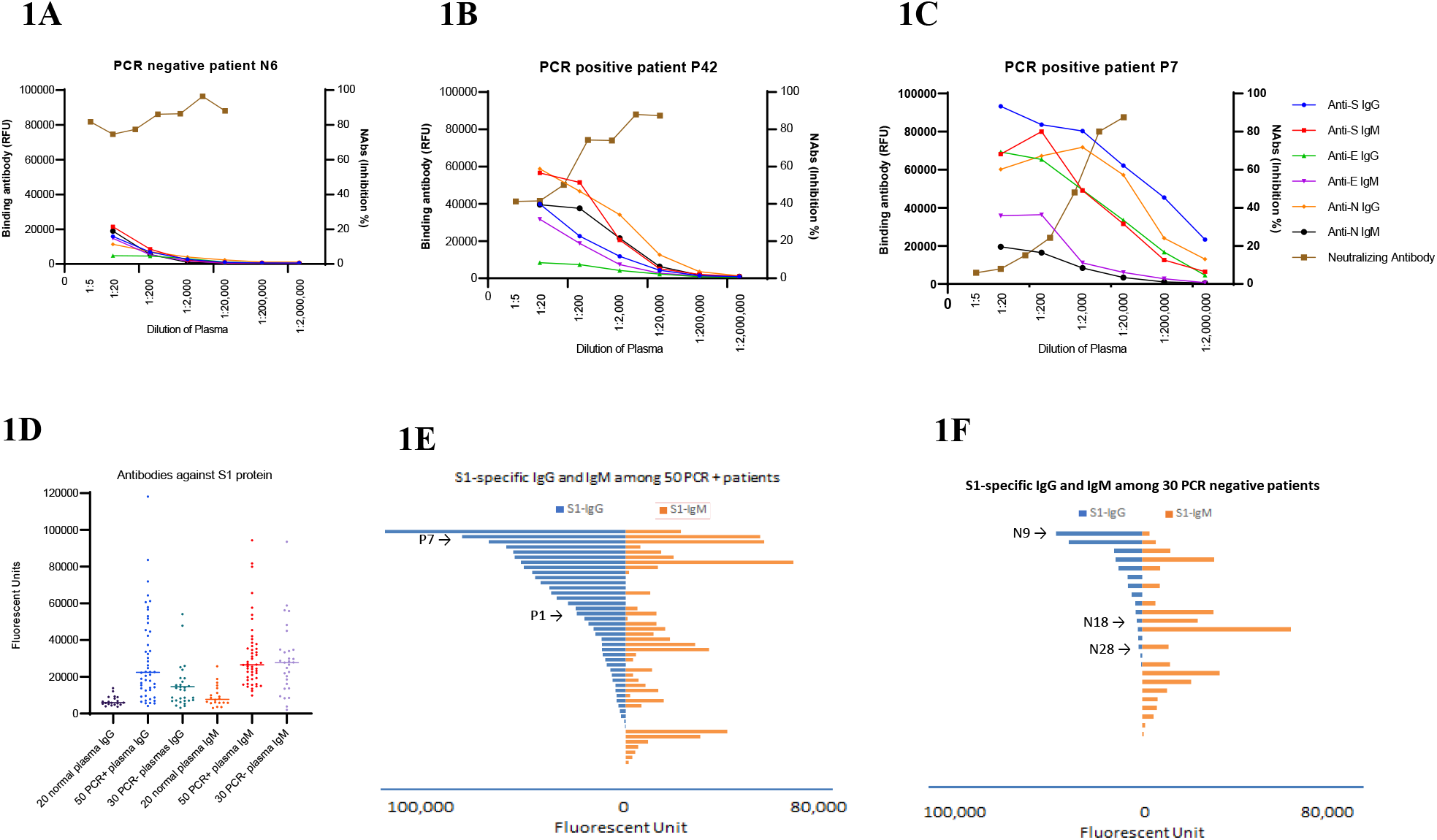
Serological evaluation of 80 plasmas from COVID-19 patients by ELISA. **A**. Antibody expression profile of PCR negative patient plasma N6 shows no SARS-Cov-2 specific binding antibody and neutralizing antibody developed; **B**. Antibody expression profile of PCR positive patient plasma P42 represents specimens with moderate level of SARS-Cov-2 antibodies; **C**. Plasma P7 profile represents a group of specimens with high level of antibody expression against S, N and E proteins and with strong neutralizing activity; **D**. Distribution of IgG and IgM against S1 protein among different specimen groups, IgG is significantly higher in PCR positive plasmas than in PCR negative plasmas but no difference was seen for IgM. **E**. Distributions of S1 IgG and S1 IgM of the 50 PCR positive patients sorted first by IgG and then IgM expression level, only displayed are the those signals above the average plus 2SD of the 20 normal plasmas; **F**. The same distributions as in Fig. 1E but for PCR negative group, the scale has been adjusted to be the same as Fig. 1E. A few representative specimens were indicated with label and arrow.

Our NAb LFT was designed to have three test lines and one control line (Fig 2A), using 60 nm Gold Nanoparticle (GNP)-conjugated RBD or S1 protein to display signal, and using ACE2 in strip (T1 line in Fig 2A) to capture GNP if the interaction between RBD and ACE2 was not completely blocked by neutralizing antibodies from a plasma. Two additional test lines showing anti-S1 protein or anti-RBD IgG (T2) and IgM (T3) were included as a positive reading reference for positive specimen.

**Figure 2.**
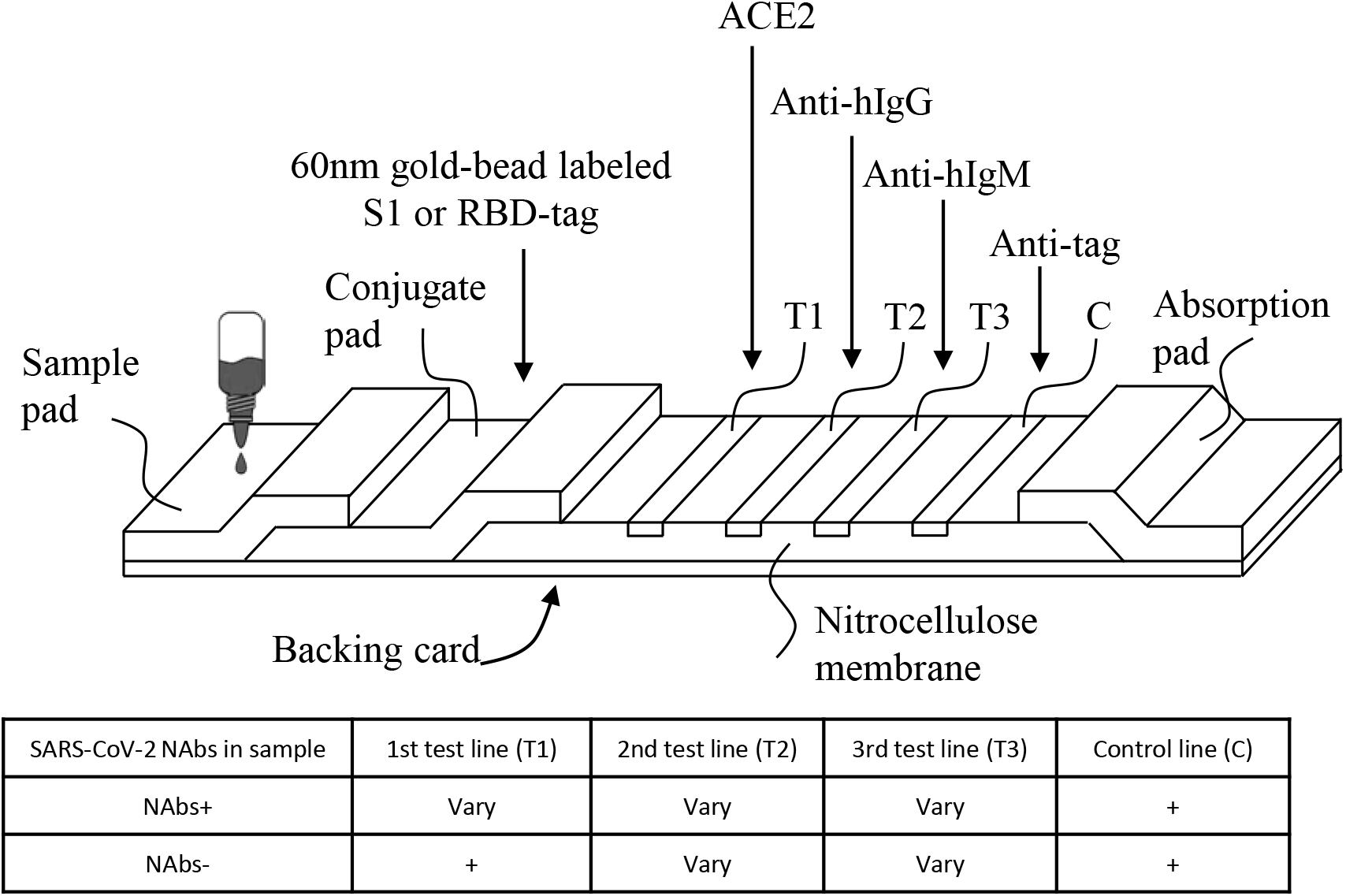
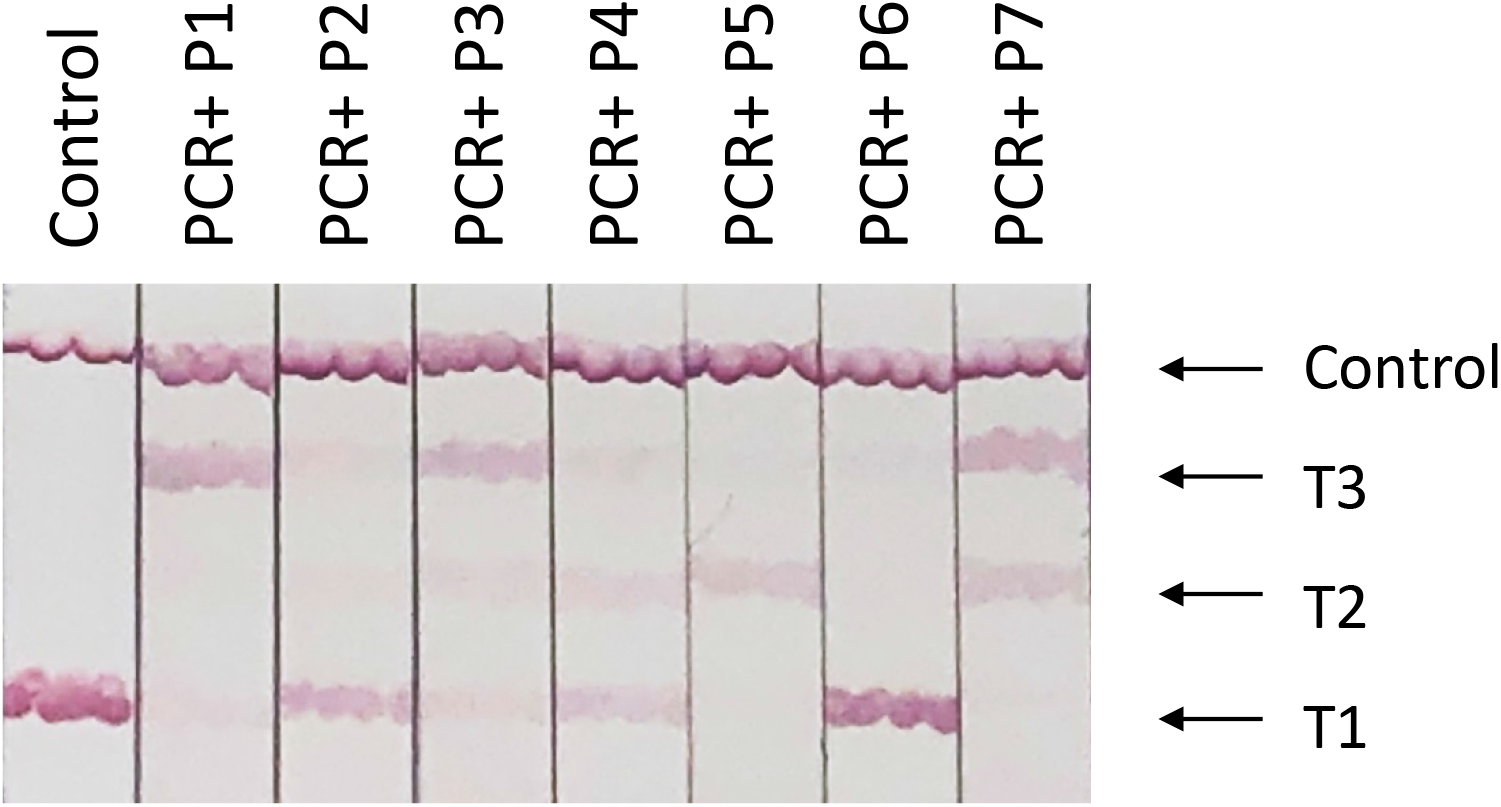
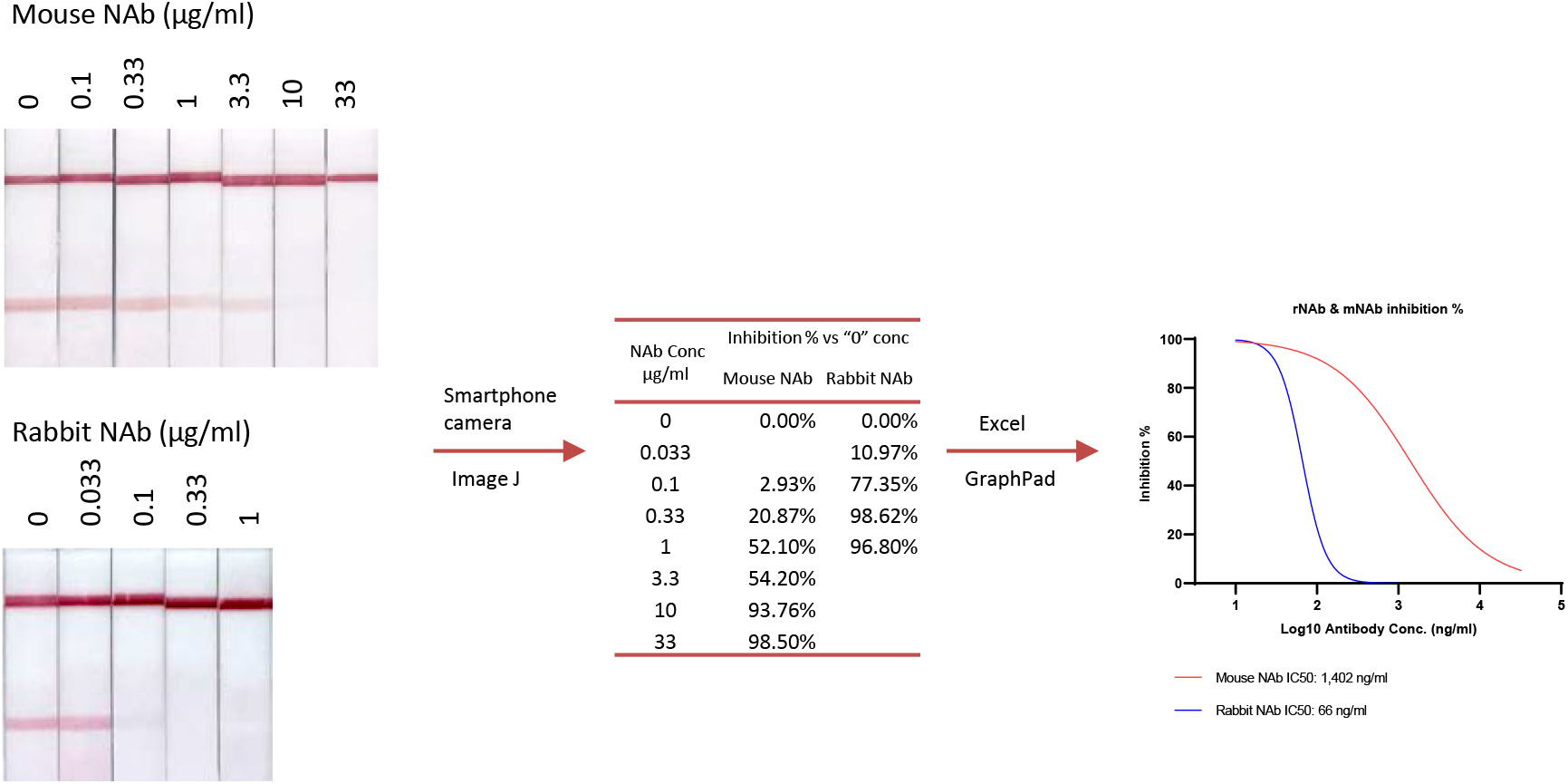
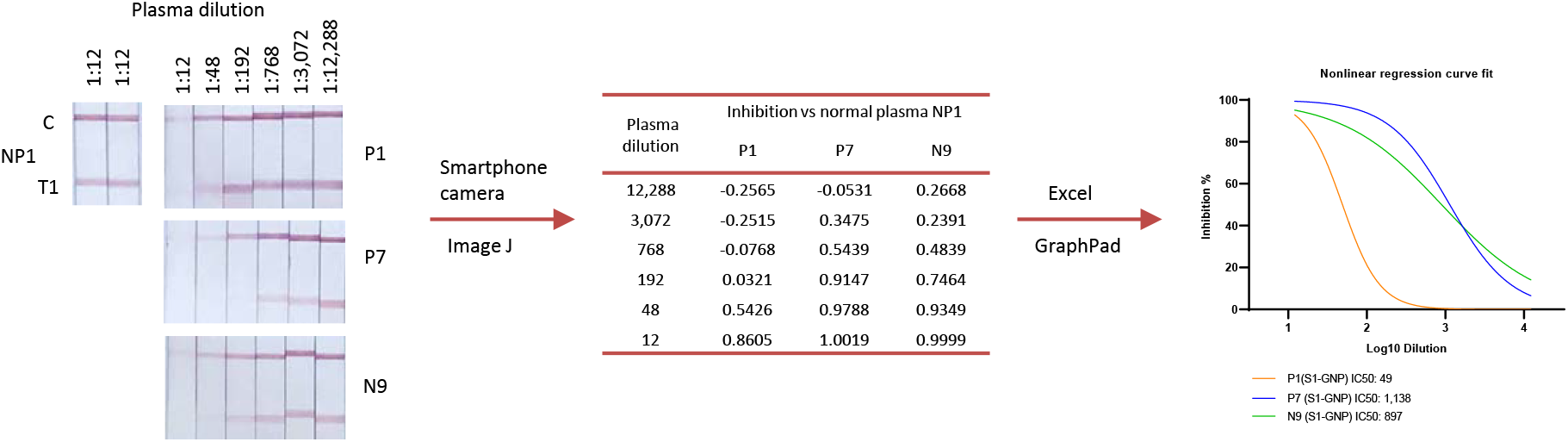
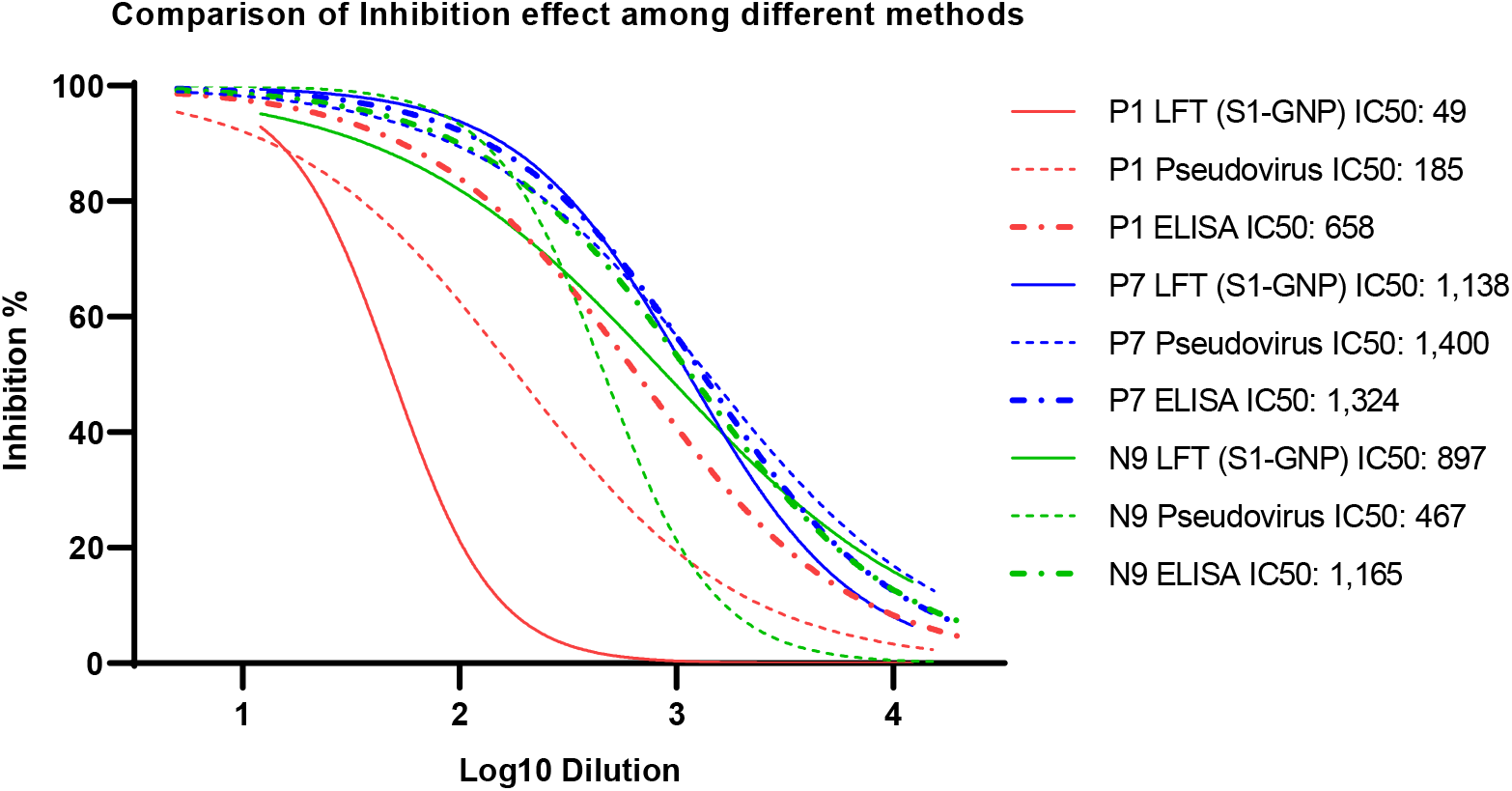
A Lateral Flow Test based on ACE2-RBD interaction. **A**. Schematic structure of the LFT strip constructed in this report and projected results. One representing format used a 60nm GNP labeled RBD with a rabbit Fc tag and accordingly an anti-rabbit IgG gamma chain antibody in control line. The strip contains three test lines, one for neutralization activity (T1) and the other two for measurement of RBD-specific IgG and IgM respectively (T2 and T3). Another 60nm GNP labeled with S1 protein was also used through the project. **B**. A manually prepared strip (using adjustable pipette to dispense/coat) demonstrating various levels of inhibition effect by different patient’s plasmas. **C**. Measurement of inhibition concentration of two neutralizing antibodies using 60nm GNP labeled RBD with a rabbit Fc tag and a workflow from 15 min test to IC50 calculation. **D**. Half-Strip demonstration of 3 plasma samples and the workflow to obtain their IC50. A GNP labeled S1 protein was used in this test. Inhibitory effects on some control lines were caused by the use of a neutralizing antibody in the control line. **E**. A comparison of three different inhibition tests, LFT, ELISA and pseudovirus based neutralization test, using one moderate and two strong inhibitory plasma specimens.

Using the above 4-line strip, we evaluated NAbs from all 80 patients’ plasmas. A single dilution of 1:12 was used for the purpose to match the common practice in the field. Typical in the field, with aid of a lancet, 10 µl of finger blood was taken and transferred into the sample pad of a LFT device, and 50-70ul of buffer was then added to drive the blood sample through the LFT device. Partial results are demonstrated in Figure 2B. The images of the test results on strips were captured with a smartphone camera and analyzed with Image J software. The intensity of the test line was computed against that of a known negative plasma and presented as inhibition percentage (Supplementary Table 1). A good correlation was observed between the inhibition percentage at 1:12 dilution of LFT and that of inhibition ELISA (r=0.79, 95% CI: 0.79-0.86, p<0.0001).

RT-PCR or PCR is currently considered as a gold standard in COVID-19 diagnosis and its result is often qualitatively presented as positive and negative. In order to parallelly compare with the qualitative PCR results, we converted all our quantitative results to qualitative positive and negative by using the cutoff established with the 20 normal plasmas for S1 IgG and S1 IgM data and by using 50% inhibition at 1:12 dilution as the cutoff for NAb measurements. A meaningful cutoff for a protective level of NAb should be established through large scale clinical trials, exemplified as IC50 of 1:10 diluted serum for Japanese Encephalitis^19^ and 1:22 for Mumps^20^. The NAb LFT cutoff we adopted here is artificial but within the reference ranges and may need to be updated later.

Correlation of our ELISA and LFT inhibition results versus patient plasma PCR results are shown in Supplementary Table 2-5. In summary, a very good negative percent agreement (90%) was observed between PCR and NAb inhibition assays among PCR negative plasmas (Supplementary Table 1, 2 and 3). The 3 plasma samples that produced discordant results (negative PCR, but positive NAb inhibition assay) have been repeatedly tested and found to contain neutralizing antibody by both inhibition ELISA and NAb LFT, thus should be considered false PCR negatives. One (plasma # N9) of those three has been chosen to demonstrate application of our NAb LFT (Fig. 2D). In contrast, IgG and IgM against S1 protein showed very poor negative percent agreement with PCR and NAb data (Fig. 1F, Supplementary Table 1, 4 and 5). 53.3% of the 30 PCR negative patients showed S1 IgG and 66.7% showed S1 IgM. It is known that the S and N proteins among all seven human infecting coronaviruses share a significant amount of sequence homology and can induce cross reactive antibody responses.^21^ To determine the contributing factor to these high negative percent disagreements, we tested 12 of the 30 PCR negative plasmas against S and N proteins of the 4 common cold coronaviruses, using the same ELISA protocol we used for SARS-CoV-2 proteins. Surprisingly, a higher IgG or IgM level against S or N proteins of at least one of the 4 common cold coronaviruses was observed, compared to that of SARS-CoV-2 among all 12 of the plasmas that were tested. Representative expression profiles by number 1 and 2 PCR negative plasmas (N1 and N2) are shown in Supplementary Figures 1A to 1D. Based on all of the available data, we believe that a certain percentage of PCR negative results were true false negatives and may be improved by testing patients’ neutralizing antibody rather than their binding antibody. Still, we cannot rule out the possibility that some of the patients in this group actually suffered severe infection caused by one of the 4 common cold coronaviruses rather than by SARS-CoV-2.

Using the strip with Positive Control and T1 lines, we demonstrated IC50 measurement of two monoclonal NAbs and three plasmas. In this experiment, a series of dilutions of each NAb or plasma were tested. The results were computed with Image J software and then analyzed and graphed with GraphPad Prism (Fig 2C and 2D). We also used the same set of patients’ plasmas to compare this procedure with inhibition ELISA and a pseudovirus neutralization test (Fig 2E). In general, we observed quite comparable IC50 results. Among the three methods, the pseudovirus neutralization test is the most complicated, expensive and lengthy. We used a commercially available pseudovirus and a one-step luciferase detection system, but still needed days of cell culture work before and after ACE2 gene transfection. Inhibition ELISA, on the other hand, has been a routine and standard method in our laboratory, like many other laboratories. It therefore provides a straightforward technical process. But even so, it requires several hours of experimental work and needs to be performed in a laboratory setting. In contrast, the NAb LFT that we’ve established works in the same way as most other lateral flow tests and can be completed within 15 minutes. Besides its convenience and short turnaround time, though, its most prominent advantage might be its nature of flexibility. In other neutralization tests, a negative specimen has to be tested for full dilution range in duplicate or triplicate in order to know the result. Using our LFT, we were able to eliminate all negative specimens after testing a single concentration and then continuously testing specimens with a higher neutralizing antibody titer until their IC50s were captured.

Theoretically, any neutralization assays using live cell and live virus simultaneously measure neutralizing antibodies targeting three regions, N terminal domain (NTD) and RBD in S1and cell fusion domain in S2. LFT and ELISA, without live cell involved and depending on which protein fragment is used, can be used to evaluate NAbs specific to one or two domains in S1 protein. We compared performance of GNP labeled RBD with GNP labeled S1 and indeed found higher NAb titer by using GNP labeled S1 (Supplementary Figure 2). However, we did not see higher NAb titer in the pseudovirus neutralization test than the other two tests in a small but direct comparison (Fig 2E).

To combat COVID-19 pandemic, our NAb LFT is continuously improved toward an IVD product. Since a neutralizing antibody result has to be quantitative or semi-quantitative, we anticipate using a smartphone and web-based solution for data capturing and processing. This not only offers convenience, but also necessary quality control for the product.

## Materials and Methods

All materials and methods are available as Supplementary information.

## Data Availability

All supplementary materials are in a single PDF file and are available online.

https://www.novodiax.com/wp-content/uploads/2020/11/A-lateral-flow-test-detecting-SARS-CoV-2-neutralizing-antibodies-Supplementary-Information.pdf

## Acknowledgments

We acknowledge the excellent proofreading and editing by David Beglin, PhD and Stephanie Wang, MD.

## Author Contributions

J.J.W. designed the NAb LFT experiments; N.Z. prepared GNP and strips; N.Z. and J.J.W. performed LFT and ELISA tests; X.Y. set-up and performed pseudovirus neutralization assays; J.V.W., S.C., N.Z. and J.J.W. contributed to data analyses; all authors contributed to the writing, editing, and completion of the manuscript.

