## Supplementary Information for "A lateral flow test detecting SARS-CoV-2 neutralizing antibodies"

**Supplementary Figure 1**: Antibody cross-reactivities among SARS-CoV-2 and common cold coronaviruses. **A.** Example demonstration using plasma N1: A comparison of IgG antibodies against S and N proteins of SARS-CoV-2 and of the 4 common cold coronaviruses using an ELISA protocol. **B.** Example demonstration using plasma N1: A comparison of IgM antibodies against the same set of proteins as shown in Supplementary Fig. 1A. **C**. Example demonstration using plasma N2: A comparison of IgG antibodies against S and N proteins of SARS-CoV-2 and of the 4 common cold coronaviruses using an ELISA protocol. **D.** Example demonstration using plasma N2: A comparison of IgM antibodies against the same set of proteins as shown in Supplementary Fig. 1C.

**Supplementary Figure 2:** A comparison of inhibition effect between GNP labeled RBD and GNP labeled S1 protein on NAb LFT. The S1 provided better inhibition in our NAb LFT strip probably because it contains two domains that can contribute to neutralization effect, RBD and NTD.

### Supplementary Figure 1A

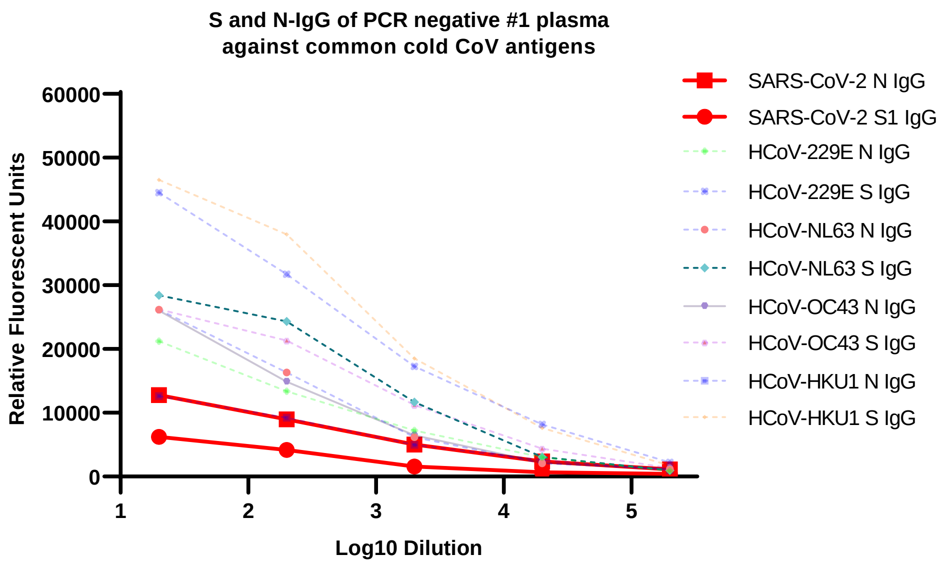


### Supplementary Figure 1B

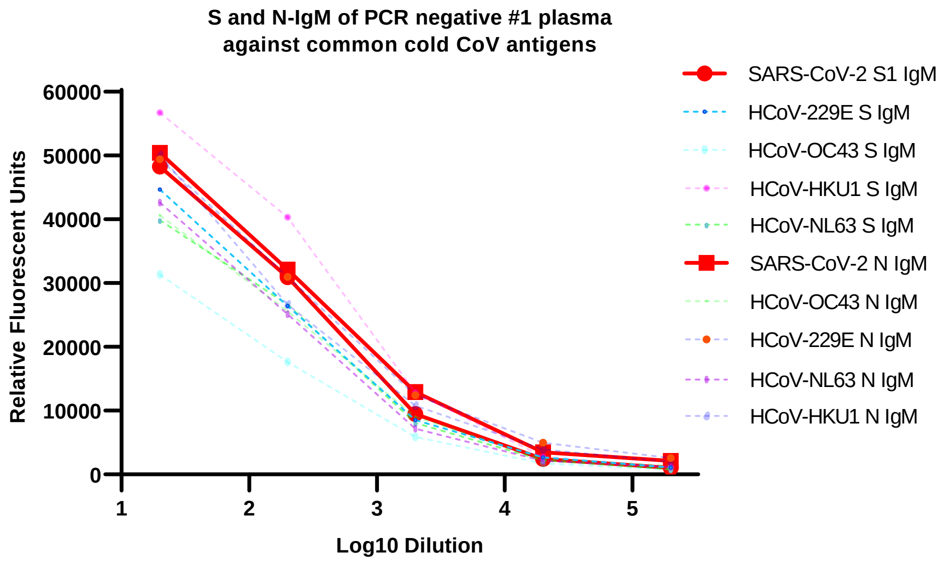

### Supplementary Figure 1C

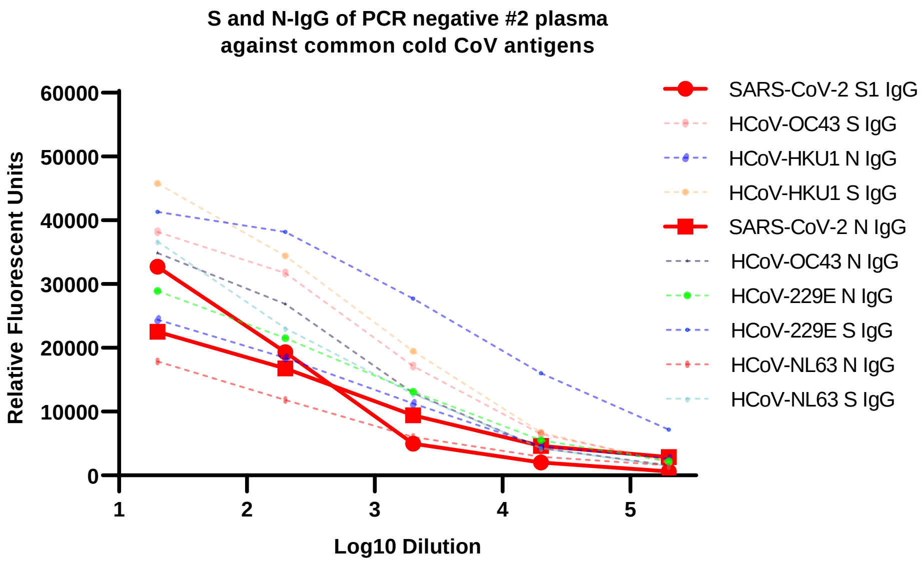


### Supplementary Figure 1D

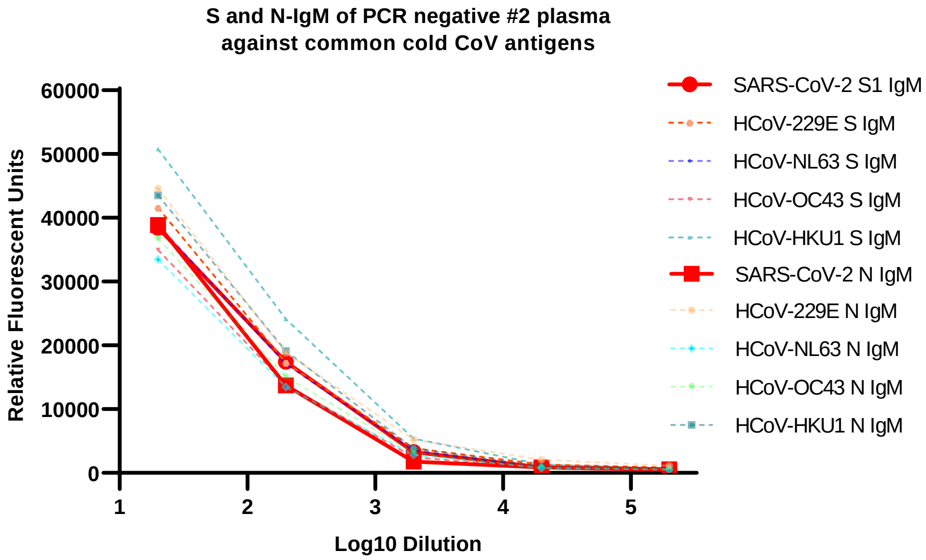

### Supplementary Figure 2

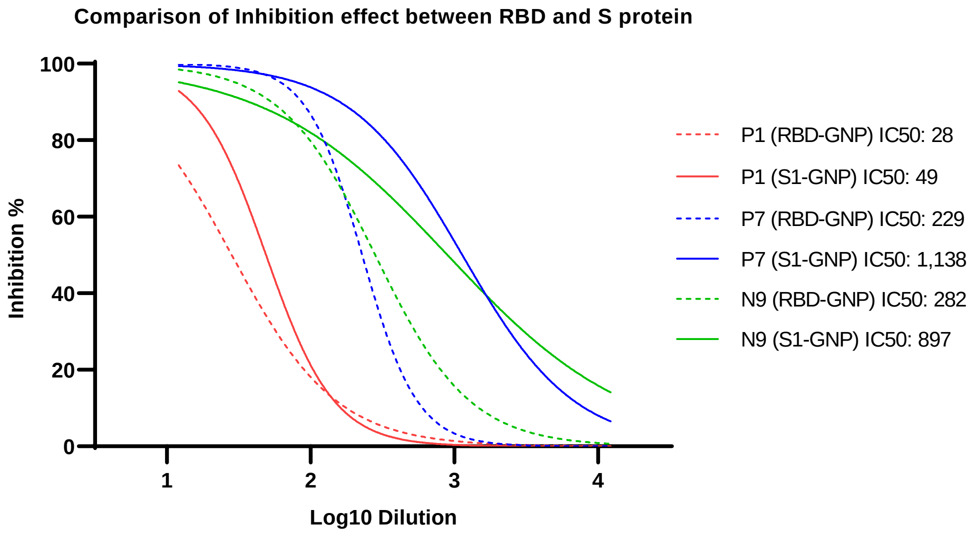

Supplementary Table 1: LFT and ELISA results of PCR positive and negative patients

| Plasma * | RT-PCR result** | S1-IgG by ELISA*** | S1-IgM by ELISA*** | Inhibition % by LFT at 1:12§ | IC50 by ELISA§§ | Inhibition % by ELISA at 1:12§§§ |
|---|---|---|---|---|---|---|
| P1 | + | 21,426 | 0 | 87 | 476 | 97 |
| P2 | + | 30,170 | 0 | 87 | 629 | 94 |
| P3 | + | 40,882 | 1,462 | 100 | 575 | 97 |
| P4 | + | 48,512 | 20,876 | 95 | 1047 | 97 |
| P5 | + | 52,131 | 6,294 | 94 | 677 | 98 |
| P6 | + | 0 | 44,310 | 0 | 25 | 60 |
| P7 | + | 71,449 | 58,667 | 97 | 1359 | 99 |
| P8 | + | 49,028 | 15,453 | 96 | 2967 | 99 |
| P9 | + | 10,537 | 19,238 | 68 | 704 | 97 |
| P10 | + | 16,289 | 13,316 | 0 | 524 | 88 |
| P12 | + | 8,923 | 3,221 | 89 | 192 | 90 |
| P13 | + | 14,090 | 17,198 | 59 | 96 | 79 |
| P14 | + | 0 | 9,711 | 0 | 27 | 57 |
| P15 | + | 6,761 | 11,498 | 0 | 58 | 65 |
| P16 | + | 0 | 4,166 | 0 | 0 | 0 |
| P17 | + | 44,479 | 13,978 | 100 | 4859 | 98 |
| P18 | + | 0 | 1,301 | 0 | 0 | 0 |
| P19 | + | 0 | 0 | 0 | 0 | 0 |
| P20 | + | 21,866 | 5,108 | 0 | 0 | 0 |
| P21 | + | 0 | 32,470 | 91 | 1207 | 88 |
| P22 | + | 6,111 | 3,136 | 80 | 1271 | 78 |
| P23 | + | 0 | 2,887 | 0 | 0 | 0 |
| P24 | + | 45,719 | 73,084 | 97 | 7282 | 98 |
| P25 | + | 3,943 | 16,502 | 0 | 1082 | 75 |
| P26 | + | 4,416 | 14,107 | 0 | 27 | 54 |
| P27 | + | 18,084 | 754 | 78 | 17 | 54 |
| P28 | + | 0 | 0 | 0 | 43 | 60 |
| P29 | + | 3,295 | 6,816 | 0 | 0 | 0 |
| P30 | + | 0 | 5,345 | 0 | 0 | 0 |
| P31 | + | 13,314 | 12,015 | 0 | 42 | 58 |
| P32 | + | 10,427 | 36,324 | 75 | 170 | 78 |
| P33 | + | 707 | 0 | 0 | 13 | 52 |
| P34 | + | 59,752 | 60,419 | 100 | 350 | 92 |
| P35 | + | 0 | 0 | 65 | 463 | 94 |
| P36 | + | 323 | 0 | 0 | 0 | 0 |
| P37 | + | 2,533 | 0 | 0 | 0 | 0 |
| P38 | + | 4,031 | 1,940 | 0 | 13 | 51 |
| P39 | + | 0 | 0 | 0 | 0 | 0 |

| | | | | | | |
|---|---|---|---|---|---|---|
| P40 | + | 5,698 | 5,590 | 0 | 18 | 56 |
| P41 | + | 10,113 | 4,395 | 66 | 651 | 94 |
| P42 | + | 10,438 | 30,214 | 0 | 61 | 71 |
| P43 | + | 33,267 | 0 | 98 | 0 | 0 |
| P44 | + | 105,943 | 24,086 | 65 | 461 | 96 |
| P45 | + | 4,481 | 8,557 | 52 | 63 | 73 |
| P46 | + | 37,181 | 0 | 68 | 53 | 96 |
| P47 | + | 39,567 | 0 | 65 | 285 | 98 |
| P48 | + | 25,165 | 0 | 0 | 0 | 0 |
| P49 | + | 8,365 | 0 | 0 | 0 | 0 |
| P50 | + | 2,120 | 0 | 0 | 0 | 0 |
| N1 | - | 0 | 7,385 | 0 | 0 | 0 |
| N2 | - | 7,119 | 0 | 0 | 0 | 0 |
| N3 | - | 0 | 7,098 | 0 | 0 | 0 |
| N4 | - | 0 | 37,513 | 0 | 0 | 0 |
| N5 | - | 0 | 5,372 | 0 | 0 | 0 |
| N6 | - | 0 | 0 | 0 | 0 | 0 |
| N7 | - | 862 | 0 | 0 | 0 | 0 |
| N8 | - | 1,986 | 0 | 0 | 0 | 0 |
| N9 | - | 41,995 | 3,513 | 100 | 2526 | 97 |
| N10 | - | 13,109 | 34,894 | 0 | 0 | 0 |
| N11 | - | 0 | 12,071 | 0 | 0 | 0 |
| N12 | - | 3,306 | 34,547 | 0 | 0 | 0 |
| N13 | - | 35,686 | 6,496 | 0 | 0 | 0 |
| N14 | - | 2,216 | 72,262 | 0 | 0 | 0 |
| N15 | - | 0 | 1,368 | 0 | 0 | 0 |
| N16 | - | 7,037 | 8,458 | 0 | 0 | 0 |
| N17 | - | 0 | 656 | 0 | 0 | 0 |
| N18 | - | 2,744 | 26,951 | 51 | 64 | 75 |
| N19 | - | 0 | 0 | 0 | 0 | 0 |
| N20 | - | 11,640 | 8,601 | 0 | 0 | 0 |
| N21 | - | 5,137 | 0 | 0 | 0 | 0 |
| N22 | - | 0 | 23,686 | 0 | 0 | 0 |
| N23 | - | 0 | 0 | 0 | 0 | 0 |
| N24 | - | 13,741 | 13,529 | 0 | 0 | 0 |
| N25 | - | 3,425 | 6,409 | 0 | 0 | 0 |
| N26 | - | 0 | 0 | 0 | 0 | 0 |
| N27 | - | 591 | 13,401 | 0 | 0 | 0 |
| N28 | - | 1,786 | 12,757 | 54 | 18 | 60 |
| N29 | - | 0 | 0 | 0 | 0 | 0 |
| N30 | - | 0 | 0 | 0 | 0 | 0 |

Supplementary Table 1 notes:

*: One of the plasmas (P11) from a PCR positive patient showed severe hemolysis, which led to high background in the strip. This plasma had to be removed from data analyses.

**: RT-PCR was performed by hospitals before the plasma specimens were collected by the Bio-Bank company iSpecimens. Details are not available.

***: ELISA results are measured in Relative Fluorescent Unit (RFU) after deduction of average RFU plus 2SD of the 20 normal plasmas.

§: LFT results are the inhibition percentage over NP1 plasma at 1:12 dilution. Less than 50% inhibition is presented as zero.

§§: Inhibition ELISA results are IC50, and all IC50 titers less than 12 are shown as zero.

§§§: IC50 generated by Inhibition ELISA was converted to inhibition percentage at 1:12 dilution using the equation Y=100/ (1+10^ ((Log10(IC50)-Log10(12)) * Hill coefficient)).

Supplementary Table 2: Agreement between RT-PCR and NAb inhibition % with 1:12 diluted plasma by LFT

| | RT-PCR + | RT-PCR - | Total |
|---|---|---|---|
| LFT NAb + | 24 | 3 | 25 |
| LFT NAb - | 25 | 27 | 54 |
| Total | 49 | 30 | 79 |

| | |
|---|---|
| Overall percent agreement | 64.4% |
| Positive percent agreement | 49.0% |
| Negative percent agreement | 90.0% |

Supplementary Table 3: Agreement between RT-PCR and NAb inhibition % with 1:12 diluted plasma by ELISA

| | RT-PCR + | RT-PCR - | Total |
|---|---|---|---|
| ELISA NAb + | 35 | 3 | 38 |
| ELISA NAb - | 15 | 27 | 42 |
| Total | 50 | 30 | 80 |

| | |
|---|---|
| Overall percent agreement | 77.5% |
| Positive percent agreement | 70.0% |
| Negative percent agreement | 90.0% |

Supplementary Table 4: Agreement between RT-PCR and S1-IgG by ELISA

| | RT-PCR + | RT-PCR - | Total |
|---|---|---|---|
| ELISA S1 IgG + | 39 | 16 | 55 |
| ELISA S1 IgG - | 11 | 14 | 25 |
| Total | 50 | 30 | 80 |

| | |
|---|---|
| Overall percent agreement | 66.3% |
| Positive percent agreement | 78.0% |
| Negative percent agreement | 46.7% |

Supplementary Table 5: Agreement between RT-PCR and S1-IgM by ELISA

| | RT-PCR + | RT-PCR - | Total |
|---|---|---|---|
| ELISA S1 IgM + | 35 | 20 | 55 |
| ELISA S1 IgM - | 15 | 10 | 25 |
| Total | 50 | 30 | 80 |

| | |
|---|---|
| Overall percent agreement | 56.3% |
| Positive percent agreement | 70.0% |
| Negative percent agreement | 33.3% |

#### Materials and Methods

##### Plasma Samples

Eighty COVID-19 patient plasma samples including 50 PCR positive and 30 PCR negative were purchased from iSpecimens (Lexington, MA). All plasma samples were incubated for half an hour in a 65℃ water-bath followed by a 5 minutes centrifuge. Supernatants were collected and 0.01% thimerosal was added as a preservative before aliquoting and storage. Twenty plasma samples collected between 2014 and 2016 before the COVID-19 pandemic were used as controls during this study.

##### Synthesis of target protein conjugated with gold nanoparticles (GNP)

The 60nm gold nanoparticle (GNP) was purchased from SigmaAldrich (Cat NO. 742015). The conjugation started by centrifuging 10ml of the GNP at 2000g for 20min. The pellet was suspended in 1mL 25mM borate buffer whose pH was adjusted to 8.5 by sodium hydroxide. The target protein was either recombinant SARS-CoV-2(2019-nCoV) Spike RBD-rFc Recombinant protein from Sino Biological Inc. (Cat NO.: 40592) or Spike/S1 protein (S1 subunit His tag) from Sino Biological (Cat NO.: 40591) was added into the GNP solution, stirred at 1000 rpm. After 1hr of incubation at ambient temperature, 0.4ml borate buffer containing 10% BSA was added into the GNP solution. The mixtures were incubated for another 1 hour at room temperature. The mixtures were then centrifuged at 2000g for 20min to discard unbound protein. The precipitate was re-suspended in 1ml PBS containing 1% BSA and stored at 4℃.

##### Preparations of capture antibodies

For this study, two major types of strips were prepared. One type was 2-line strips designed for neutralization sensitivity tests, and the other type was 4-line strips designed for testing the plasma samples from patients. The 2-line strips contained alpaca anti-rabbit IgG (H+L) from Jackson ImmunoResearch (Code: 611-005-215) or Rabbit anti mouse Fcϒ fragment specific from Jackson ImmunoResearch (Code: 315-005-046) at control line and ACE2 protein from Sino Biological Inc. (Cat NO.: 10108) at T1 line. For the 4-line strips, donkey anti-human IgM, Fc5μ from Jackson ImmunoResearch (Code: 709-005-073) and donkey anti human Fcϒ from Jackson ImmunoResearch (Code: 709-005-098) were used at T3 and T2, respectively. All the capture protein or antibodies were mixed with 2% trehalose and 10% sucrose before being applied onto nitrocellulose membranes. The concentration of each capture antibody was tested and optimized prior to use.

**Protocol to assemble Lateral Flow Test (LFT) strips**

Several instruments were employed for this research. First, an automated lateral flow reagent dispenser from Claremont Bio was used for applying antibodies to Whatman FF170HP membranes. The membranes were then dried at 40°C for 1 hour, blocked with PBS-1% BSA for 5 min and dried again in a 40°C oven for an hour. Then, 2cm wide wick pad and sample pad were cut by a paper trimmer. The Biodot LM5000TM Lamination system was used to assemble the membrane, sample pad, and wick pad. Lastly, a Matrix 2360 programmable shear was used to cut the assembly into 0.5cm wide strips. All the strips were refrigerated in a vacuum sealed bag for storage.

**Neutralization Test by LFT**

A half-strip format was used in most of LFT neutralization tests. A ninety-six well microtiter plate was used to hold testing solution and strips during the test. For each testing solution, 5µL of plasma or testing sample was added to a mixture of 5µL of GNP conjugated RBD or S1 recombinant protein and 50µL of PBS buffer containing EDTA. Final dilution of the plasma was 1:12. The strips were then dipped into the solution and left for 15 minutes. The results were then recorded by a smartphone camera, converted to digital signals by Image J, and calculated and graphed by GraphPad Prisma 8 software. The tests were conducted in duplicates for the generation of IC50. A normal plasma was always included as a negative control in the test. The two monoclonal neutralizing antibodies used during the development of this test were from Sino Biological Inc. (Cat NO. 40592-R001 and 40592-MM57).

**ELISA**

Recombinant SARS-CoV-2 S1, N and E proteins and S and N proteins from 4 common cold coronaviruses, all purchased from Sino Biological, were coated 25µl/well onto black high binding ELISA 384-well plates (Greiner Bio-One) at a concentration of 0.25µg/mL in 50mM carbonate buffer, pH 9.6. Then the plates were incubated overnight at 4°C. After being washed 3 times by PBS-T, PBS containing 1% BSA were applied to each well as a blocker. After one hour blocking at room temperature, plasma samples were diluted by the blocker to 1:20, 1:200, 1:2k,

1:20k and 1:200k, and were added and incubated for 1 hour at room temperature. After washing another three times, the plates were incubated with HRP-conjugated goat anti human IgG or IgM (Jackson ImmunoResearch) for another hour at room temperature. After washing five more times, Amplex Red (Thermo Fisher) was used to develop a fluorescent signal. Plates were read by Flexstation 2 (Molecular Devices).

**Inhibition ELISA**

This protocol was modified from the ELISA protocol above. The plates were coated with RBD-rFc at 0.25ug/ml for o/n at 4 deg C. Mix 1ug/ml of ACE2-mFc with serially diluted plasma (1:5, 20, 80, 400, 1600, 6400, 25600, 102400). They were then added to RBD-rFc coated plates and incubated for 1h. Next a HRP labeled anti-mouse IgG with minimal cross-reactivity with human IgG was used to generate a detection signal.

**Pseudovirus Neutralization Test**

**Cells:** HEK-293 cells (35mm dish) at approximately 85% confluence were transfected with hACE2 plasmid (Addgene plasmid #1786, 500ng plasmid + 10ul 293fectin from Thermo fisher Cat# 12347019) with 200ul Opti-DMEM; 48 hours after transfection, cells were seeded into 384 well plates at 7500 cells/well/25ul 10% FBS complete DMEM medium; Transfected HEK-293 cells were confirmed for expression of hACE2 by cytology- 95%-100% of transfected cells expression hACE2。 **Plasma neutralization assay:** Plasma was used at 1: 5 serial dilution (final concentration: 1:5, 1:25, 1:75; 1:375, 1:1875, and 1:9375); SARS-CoV-2 S lenti Pseudo viruses (BPS Bioscience Cat# 79942) were used at 500 viruses/well; Polybrene Transfection Reagent (Millipore) was used at final 1ug/ml to help the virus infection; Diluted plasma (or neutralizing Ab used at final concentration: 5ug/ml, 1 ug/ml, 0.2ug/ml, 0.04ug/ml, 0.08ug/ml, and 0.016ug/ml) and Pseudo viruses were mixed and incubated at RT for 30 min; Approximately 25ul of plasma and Pseudo viruses' mixtures were added to each well with hACE2 expressing cells (totally 50 ul) in triplicates for each condition; Spin down the plate at 1500g for 15 min to help virus infection; Incubate cells at 37$^{o}$C for 48 hours; detect luciferase activity using One-Step Luciferase Assay System (BPS Bioscience Cat #60690-1); approximately 50 ul of luciferase assay working solution (Component A + Component B) were added to each well (totally 100 ul volume); gently rock the plate for 15 min at RT and Spin down at 1000g for 3 min; measure firefly luminescence using a luminometer (Thermo-Fisher Fluoroskan-FL).

**Image acquisition and analysis**

The lateral flow strips were imaged by a mobile phone under even LED light illuminations. The images were downloaded and analyzed by ImageJ (National Institutes of Health, Bethesda, MD, USA). In brief, after image conversions, the rectangle region of interest (ROI) was selected and computed to obtain mean gray value.  Adjacent background gray value on the same strip was subtracted.  Stain intensity was then normalized to that of control to get percentage signal intensity.
